# Concordance with the MIND diet is associated with lower subsequent epigenetic aging

**DOI:** 10.64898/2026.10.30.26365745

**Authors:** Vivian Hsing-Chun Wang, Jordan Weiss, Yian Gu, Jeannette Beasley, Emily Johnston, Arjun Masurkar, Daniel Wronski, Donglan S. Zhang

## Abstract

Healthy dietary patterns may influence biological aging, but prospective evidence for the Mediterranean–DASH Intervention for Neurodegenerative Delay (MIND) diet is limited. We linked 2013 dietary data to 2016 DNA methylation measures in 1,740 Health and Retirement Study participants. Per 1-s.d. higher MIND score, adjusted differences were −0.4 years for PhenoAge acceleration (95% CI −0.8 to −0.1), −0.3 years for GrimAge acceleration (−0.5 to −0.1) and −0.0054 for methylation Pace of Aging (−0.0098 to −0.0010); all false-discovery-rate q≤0.026. Associations persisted after body-mass-index adjustment and selection weighting. Targeted maximum likelihood estimates were concordant with the conventional models, whereas causal forests evaluated in held-out and repeated-split samples found no reproducible evidence that specific subgroups of participants benefited more than others. Greater MIND concordance was associated with lower subsequent epigenetic aging across complementary measures, supporting molecular aging as an intermediate phenotype for diet-aging research.

## Introduction

A central goal of geroscience is to identify modifiable exposures that influence the biological processes linking chronological aging to chronic disease, functional decline and mortality.^1^ Diet is a plausible candidate because dietary patterns act across cardiometabolic, inflammatory and neurocognitive pathways rather than through a single organ system. Recent population studies have linked greater concordance with healthy dietary patterns with slower accumulation of multimorbidity in older adults and a higher probability of reaching later life free of major chronic disease and functional impairment.^2,3^

The Mediterranean–DASH Intervention for Neurodegenerative Delay (MIND) diet incorporates components of Mediterranean and Dietary Approaches to Stop Hypertension dietary patterns with additional emphasis on foods proposed to support brain health.^4,5^ Both patterns emphasize foods such as vegetables, berries, whole grains, nuts, beans, fish, poultry, and olive oil. These diets limit foods such as red meat, butter and margarine, cheese, pastries and sweets, and fried foods. Green leafy vegetables and berries are highlighted partly because of their relatively high content of vitamin K, folate, and flavonoids.^6,7^ In observational studies, greater concordance has been associated with slower cognitive decline, lower Alzheimer’s disease incidence, reduced β-amyloid load in Alzheimer’s disease, and decreased brain atrophy.^4,5,8,9^ Yet in a 3-year randomized trial, participants assigned to the MIND diet with mild caloric restriction showed no significant difference in cognitive change relative to those in an active control group that followed a caloric-restriction diet.^10^ Although both groups experienced a benefit in global cognition,^10^ these findings make it especially important to distinguish associations between dietary patterns and intermediate biological pathways from evidence that a MIND diet concordance improves clinical outcomes.

While the literature has demonstrated an association between cardiometabolic factors and brain aging as well as a relationship between inflammatory biomarkers and cognitive outcomes in context of MIND diet concordance,^11,12^ a secondary analysis of the Women’s Health Initiative data has shown an inverse relationship between diet-quality scores and DNA methylation-based aging measures. DNA methylation influences gene expression without altering the underlying DNA sequence,^13,14^ and changes in DNA methylation is associated with aging, neurodegenerative disease, and Alzheimer’s disease.^15^ 15 DNA-methylation epigenetic clocks, including GrimAge, Levine/PhenoAge, and the pace-of-aging measure known as the Dunedin Pace of Aging (PoAm) DNAm clock. estimate epigenetic age and can be compared alongside an individual’s chronological age as epigenetic age acceleration, capturing aspects of aging that are not fully explained by chronological age alone.^16^ PhenoAge captures methylation signatures associated with a phenotypic-age index incorporating 9 clinical biomarkers and mortality risk.^17^ GrimAge incorporates methylation surrogates of 7 mortality-associated plasma proteins and smoking exposure, strongly predicting all-cause mortality, as well as risk of coronary heart disease and cancer development.^18^ The DunedinPoAm38 methylation clock was trained on the 12-year changes in 18 blood-chemistry and organ-system-function biomarkers to estimate the rate of aging rather than age at a single point in time.^19–21^ Recent work in *Nature Aging*has further illustrated that DNA methylation measures can capture functional and lifestyle-related variation in aging not represented by chronological age alone.^22^ However, epigenetic clocks are not interchangeable measures of a single latent process, and concordance across clocks trained on different aging-related targets is therefore potentially informative.

Evidence connecting overall diet quality to epigenetic aging has expanded rapidly. In the INSPIRE-T cohort, greater a DASH pattern was associated with lower PhenoAge acceleration, although findings were not consistent across other clocks and were partly attenuated after accounting for adiposity.^23^ More recently, cross-sectional baseline analyses from the Rhineland Study, with replication in EPIC-Potsdam, showed that healthier dietary patterns generally tracked with lower PhenoAge acceleration, GrimAge acceleration and DunedinPACE; MIND concordance was directionally favorable across these measures, although the strength and reproducibility of associations varied across dietary scores.^24^ These studies establish diet quality as a correlate of molecular aging but leave uncertainty about temporal ordering and about whether a brain-health-focused diet predicts later epigenetic aging in a population-based US cohort.

To address this gap, we linked a 164-item food frequency questionnaire from the 2013 Health Care and Nutrition Study capturing intake within the previous 12 months to DNA methylation measures collected approximately three years later in the Health and Retirement Study (HRS). We examined associations of MIND diet concordance with PhenoAge acceleration, GrimAge acceleration and methylation Pace of Aging, tested robustness to socioeconomic and behavioral confounding, adiposity and selection into the methylation sample, and explored whether any association was concentrated among different population groups. We hypothesized that greater MIND concordance would be associated with lower subsequent epigenetic aging across complementary measures.

## Results

### Study population and MIND diet concordance

Among 7,608 HRS participants with a derived MIND score^5^ and energy-intake data from the 2013 Health Care and Nutrition Study, 1,754 matched to the 2016 epigenetic-clock data before applying energy-intake exclusions. After exclusions were applied in the nutrition source cohort and the analysis was restricted to participants with DNA methylation data, 1,740 participants comprised the descriptive analytic sample (Flowchart shown in Fig. 1). The primary multivariable models included 1,722 complete cases.

**Fig. 1.**
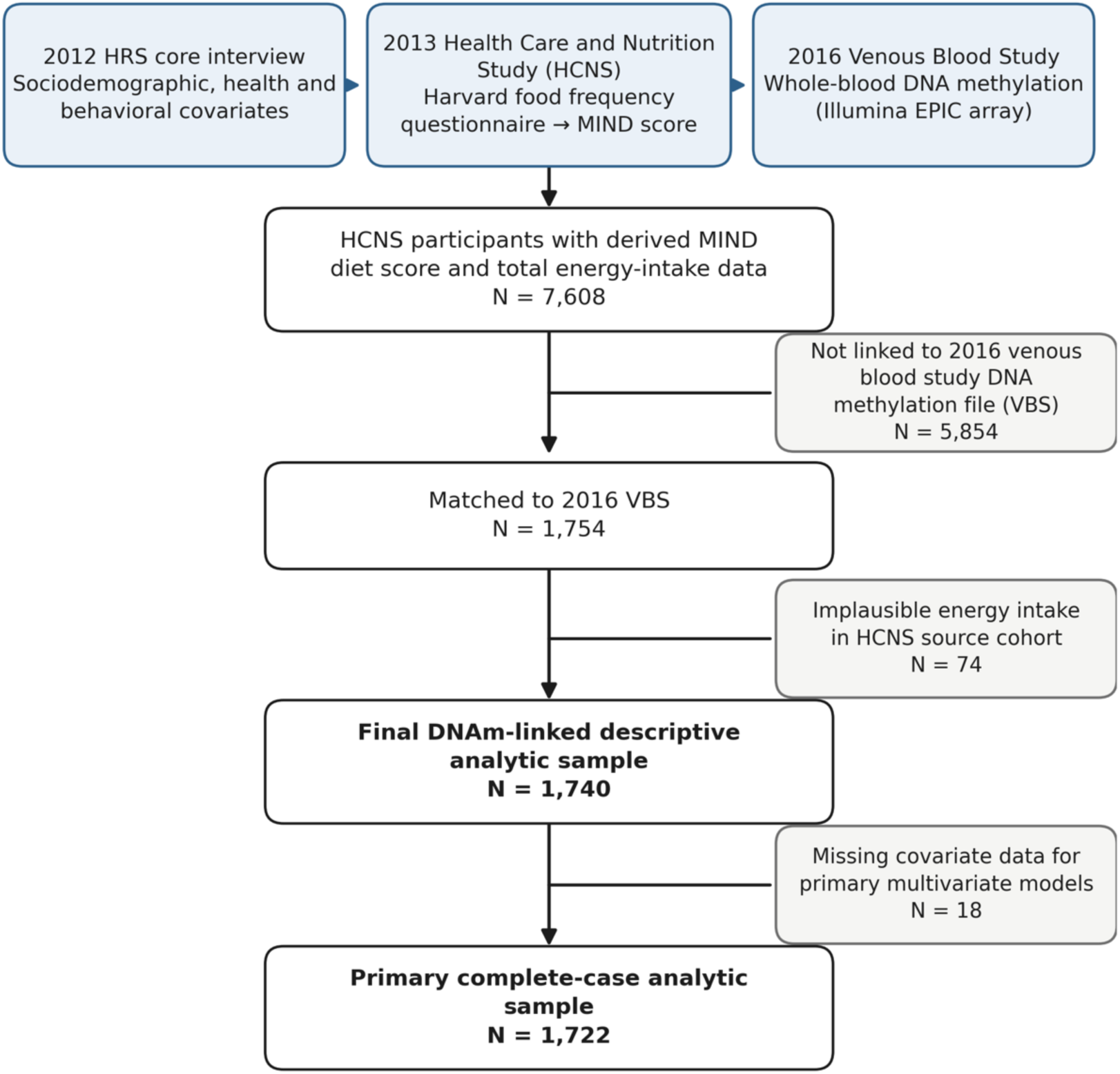
Study design and analytic flow. Timeline and participant flow linking covariates from the 2012 HRS core interview, dietary assessment from the 2013 Health Care and Nutrition Study and DNA methylation from the 2016 Venous Blood Study. The HCNS source cohort included 7,608 participants with derived MIND score and energy-intake data; 1,754 matched to the epigenetic-clock file before dietary energy exclusions, and 1,740 comprised the final DNA methylation descriptive sample after exclusions were applied in the source cohort. Primary complete-case models included 1,722 participants.

Participants ranged from 47 to 93 years of age and had a mean (s.d.) age of 66.0 (9.5) years; 1,033 (59.4%) were women. The mean (s.d.) MIND score was 7.2 (1.9) out of a maximum possible score of 15. MIND diet concordance was measured by MIND score tertiles, with higher tertile showing better concordance. Participants in the highest MIND tertile had a mean score of 9.5 compared with 5.4 in the lowest tertile. Greater MIND concordance was descriptively associated with higher educational attainment and household wealth, lower current smoking, greater physical activity and lower mean body mass index (Extended Data Table 1).

### Greater MIND concordance is associated with lower epigenetic aging

In primary models adjusted for age, sex, race and ethnicity, educational attainment, household wealth, marital or partner status, smoking history, physical activity and total energy intake, greater MIND concordance was inversely associated with all three epigenetic-aging measures (Fig. 2). Each 1-s.d. higher MIND score, corresponding to approximately 1.9 points on the 15-point scale, was associated with 0.404 fewer years of PhenoAge acceleration (95% CI −0.758 to −0.049; false-discovery-rate q=0.026), 0.309 fewer years of GrimAge acceleration (95% CI −0.502 to −0.116; q=0.005) and a 0.0054-unit lower methylation Pace of Aging (95% CI −0.0098 to −0.0010; q=0.023). The direction of association was therefore consistent across a phenotypic-age clock, a mortality-trained clock and a rate-like measure of aging.

**Fig. 2.**
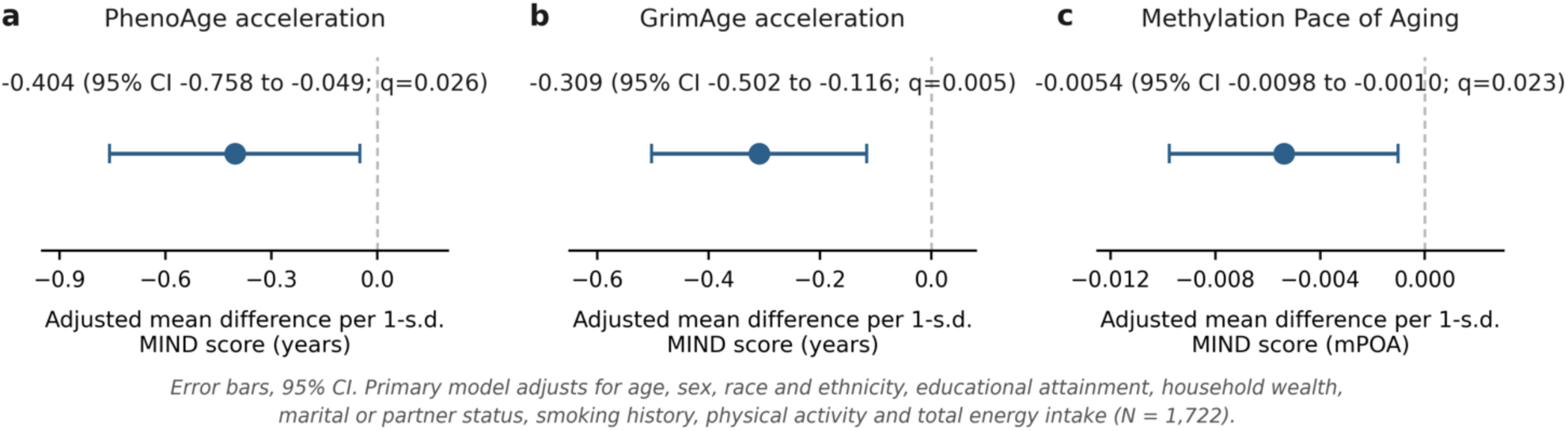
MIND diet concordance and subsequent epigenetic aging. Adjusted mean differences (points) and 95% confidence intervals (error bars) in PhenoAge acceleration, GrimAge acceleration and methylation Pace of Aging per 1-s.d. higher MIND score, from the primary model adjusting for age, sex, race and ethnicity, educational attainment, household wealth, marital or partner status, smoking history, physical activity and total energy intake (N = 1,722). q values reflect the Benjamini-Hochberg false-discovery-rate procedure applied across the three outcomes. Negative values indicate lower (slower) epigenetic aging with higher MIND concordance.

Associations were larger in minimally adjusted models and attenuated after addition of socioeconomic and behavioral covariates, particularly for GrimAge and methylation Pace of Aging (Extended Data Table 2). Additional adjustment for body mass index produced only modest further attenuation. The corresponding estimates were −0.378 years for PhenoAge acceleration (95% CI −0.733 to −0.022), −0.287 years for GrimAge acceleration (95% CI −0.478 to −0.095) and −0.0050 for methylation Pace of Aging (95% CI −0.009 to −0.001).

Secondary analyses using MIND tertiles showed a generally graded pattern in adjusted outcome means (Fig. 3). Relative to the lowest tertile, the highest tertile had lower GrimAge acceleration (−0.675 years, 95% CI −1.110 to −0.241) and lower methylation Pace of Aging (−0.012, 95% CI −0.022 to −0.002). PhenoAge acceleration was also lower in the highest MIND tertile (−0.777 years), although its 95% CI included the null (−1.597 to 0.044). Estimates comparing the middle with the lowest tertile were smaller across all three outcomes.

**Fig. 3.**
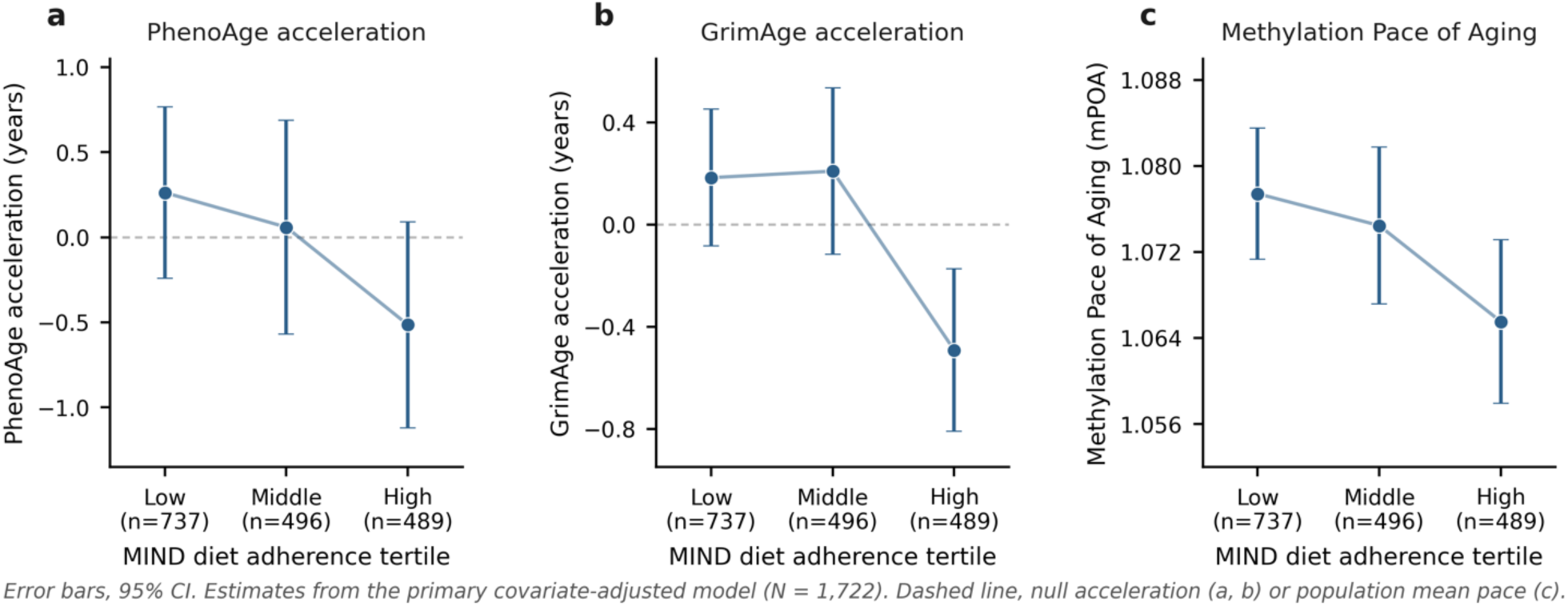
Adjusted epigenetic-aging measures across MIND diet concordance tertiles. Covariate-adjusted means and 95% confidence intervals for PhenoAge acceleration, GrimAge acceleration and methylation Pace of Aging across low, middle and high MIND-concordance tertiles, from the primary multivariable model adjusting for age, sex, race and ethnicity, educational attainment, household wealth, marital or partner status, smoking history, physical activity and total energy intake (N = 1,722). Dashed reference lines mark the null (no acceleration, PhenoAge/GrimAge) or the reference pace of one biological year per chronological year (1.0, methylation Pace of Aging). Lower values indicate slower epigenetic aging.

### Associations persist after selection weighting and do not differ by age or sex

There was no evidence that the continuous association of MIND concordance with any epigenetic-aging measure differed by sex or chronological age. MIND-by-sex interaction P values were 0.454 for PhenoAge acceleration, 0.860 for GrimAge acceleration and 0.904 for methylation Pace of Aging; corresponding MIND-by-age interaction P values were 0.525, 0.940 and 0.802, respectively (Extended Data Fig. 1 and Supplementary Table 1).

We next addressed measured selection from the 2013 nutrition cohort into the later DNA methylation sample. The selection model incorporated MIND score, age, sex, race and ethnicity, education, household wealth, marital status, smoking, physical activity, energy intake and body mass index and combined the inverse predicted probability of DNA methylation inclusion with the Health Care and Nutrition Study base weight. In selection-weighted models (n=1,644), each 1-s.d. higher MIND score was associated with −0.714 years of PhenoAge acceleration (95% CI −1.177 to −0.250), −0.376 years of GrimAge acceleration (95% CI −0.604 to −0.148) and −0.0082 units of methylation Pace of Aging (95% CI −0.013 to −0.003) (Extended Data Table 3). Thus, accounting for measured differences between the nutrition source cohort and methylation subsample did not attenuate the primary associations.

### Doubly robust estimates support an average association but not reproducible heterogeneity

We used exploratory targeted-learning analyses to evaluate a binary contrast between participants in the highest MIND tertile and those in the lower two tertiles. Among 1,722 complete cases, 489 were in the highest tertile and 1,233 were in the lower two tertiles. Targeted maximum likelihood estimation yielded average differences of −0.894 years for PhenoAge acceleration (95% CI −1.663 to −0.126; q=0.023), −0.847 years for GrimAge acceleration (95% CI −1.268 to −0.426; q<0.001) and −0.0125 for methylation Pace of Aging (95% CI −0.0223 to −0.0027; q=0.015) (Fig. 4 and Extended Data Table 4). Overlap-targeted augmented inverse-probability-weighted (AIPW) estimates were similar, as were targeted maximum likelihood estimates after additional body-mass-index adjustment. A sensitivity contrast restricted to the highest and lowest MIND tertiles likewise produced estimates in the same direction for all three outcomes.

**Fig. 4.**
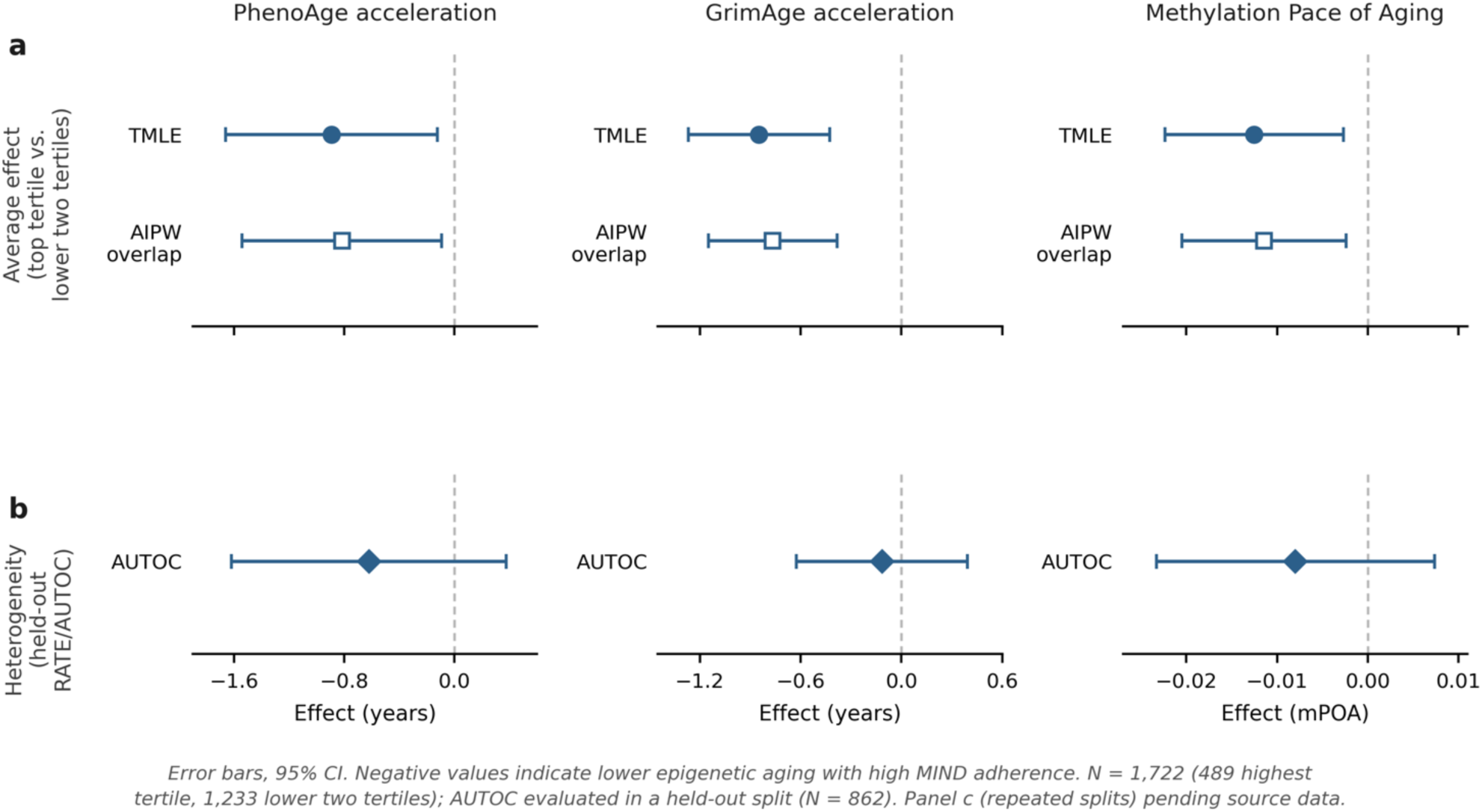
Average and heterogeneous associations of high MIND concordance with epigenetic aging a, Targeted maximum likelihood and overlap-targeted augmented inverse-probability-weighted estimates comparing the highest MIND tertile with the lower two tertiles (N = 1,722; 489 highest tertile, 1,233 lower two tertiles). Negative original-scale values indicate lower epigenetic aging with high MIND concordance. **b,** Held-out rank-weighted average treatment effect (AUTOC) estimates testing for treatment-effect heterogeneity in the main train-evaluation split (evaluation N = 862). **c,** Median AUTOC (interquartile range) from the same held-out heterogeneity test repeated across five additional random splits. The analyses support average associations across all three aging measures but do not identify reproducible heterogeneity.

In contrast, there was no evidence that the average association was concentrated among a reproducible subset of participants. In held-out evaluation samples, rank-weighted average treatment effect analyses yielded AUTOC estimates whose 95% CIs included the null for PhenoAge acceleration (−0.621, 95% CI −1.620 to 0.377), GrimAge acceleration (−0.115, 95% CI −0.624 to 0.394) and methylation Pace of Aging (−0.0080, 95% CI −0.0233 to 0.0073) for the primary high-versus-lower contrast; all false-discovery-rate q values were at least 0.647. The highest-versus-lowest-tertile sensitivity contrast was also null across outcomes. Direct comparisons between the top and bottom 20% of predicted benefit were not significant after false-discovery-rate correction.

The absence of stable heterogeneity was reinforced by repeated held-out analyses. Across five additional random train–evaluation splits, none of the primary-contrast AUTOC tests was nominally significant for any outcome. Median AUTOC values were −0.360 for PhenoAge acceleration, −0.003 for GrimAge acceleration and −0.003 for methylation Pace of Aging, and the direction of the predicted ranking was not stable across splits for GrimAge or methylation Pace of Aging (Supplementary Table 2). Together, these analyses supported an average association between MIND concordance and lower epigenetic aging but provided little evidence for a robust precision-nutrition subgroup effect.

## Discussion

In this population-based study of middle-aged and older US adults, greater concordance with the MIND diet was associated with lower epigenetic aging measured approximately three years later. The association persisted after adjustment for demographic characteristics, educational attainment, household wealth, smoking history, physical activity and energy intake and was observed across PhenoAge acceleration, GrimAge acceleration and methylation Pace of Aging. All three primary associations remained after false-discovery-rate correction, were only modestly attenuated by additional body-mass-index adjustment, and were maintained in selection-weighted analyses. Exploratory targeted-learning estimates were concordant with the conventional models, whereas held-out causal forests provided no reproducible evidence that the association was concentrated among specific participant profiles.

The findings add temporal information to a growing evidence base that has expanded substantially over the past year. In the largest of such analyses to date, spanning ten diet-quality scores across the recent Rhineland Study and an independent EPIC-Potsdam replication, healthy dietary patterns, generally tracked with lower PhenoAge acceleration, GrimAge acceleration and DunedinPACE, although MIND concordance specifically showed its most consistent, false-discovery-rate-corrected association with the pace-of-aging measure only.^24^ That study used cross-sectional baseline diet and methylation measurements in its primary cohort.^24^ INSPIRE-T similarly reported an inverse association of DASH concordance with PhenoAge acceleration, but not consistent associations with GrimAge or other aging measures.^23^ Our results extend these observations by measuring diet approximately three years before DNA methylation and by focusing on a US cohort spanning midlife through advanced age. Our findings also complement recent *Nature Aging* evidence that healthy dietary patterns are associated with slower accumulation of multimorbidity in older adults.^2^ Taken together, these studies suggest that diet quality may relate both to molecular aging phenotypes and to the downstream accumulation of age-related disease, although the observational evidence does not establish that these relationships form a single causal pathway.

The consistency of direction across epigenetic measures is notable because the clocks capture related but nonidentical constructs. PhenoAge was developed to predict a clinically derived phenotypic age and mortality, whereas GrimAge was trained more directly on mortality-related plasma-protein surrogates and smoking history.^19,25^ DunedinPoAm38 is conceptually different again, estimating a pace of physiological deterioration derived from longitudinal changes across organ systems.^21^ The three measures are therefore not independent replications, but concordant findings across them reduce the likelihood that the association reflects a single clock-specific architecture. Recent work in *Nature Aging* has similarly emphasized that aging biomarkers can capture distinct functional and lifestyle-related dimensions of aging rather than a unitary biological age.^22^

Adjustment for socioeconomic and behavioral factors materially attenuated associations relative to models containing only age, sex, and race and ethnicity, underscoring the extent to which diet quality is embedded within broader social and behavioral environments. This was especially apparent for GrimAge and methylation Pace of Aging. Household wealth is relevant in HRS because socioeconomic gradients are strongly associated with second- and third-generation epigenetic clocks, including GrimAge and DunedinPoAm.^26^ Smoking history is particularly important for GrimAge because the clock incorporates a methylation surrogate of cumulative smoking exposure.^25^ Even after explicitly controlling for these factors, however, the MIND association remained. Prior HRS work has also linked physical activity with lower epigenetic aging, reinforcing the importance of treating healthy behaviors as correlated exposures rather than isolated interventions.^27^

Several biological pathways could plausibly contribute to the observed association. The MIND diet emphasizes vegetables, berries, legumes, nuts, whole grains, fish, poultry and olive oil while limiting red and processed meat, butter, cheese, sweets and fried foods, and alcohol intake. These food categories and characteristics may influence inflammation, oxidative stress, insulin sensitivity, lipid metabolism, endothelial function, the gut microbiome and adiposity, all of which are implicated in age-related molecular dysregulation.^28–30^ The recent epigenome-wide diet analysis found that distinct healthy dietary patterns mapped to partially distinct Cytosine-phosphate-Guanine (CpGs)—where DNA methylation mainly occurs—but converged strongly on overlapping biological pathways, with MIND-associated methylation signals particularly implicating cognition-, learning- and axonogenesis-related processes.^24^ Our modest attenuation after body-mass-index adjustment is compatible with adiposity contributing to, but not fully accounting for, the association. Because long-term dietary patterns can influence body weight, BMI may lie in the causal pathway between diet and epigenetic aging and therefore act as a mediator rather than solely a confounder. The BMI-adjusted models should not, for that reason, be interpreted as inherently less confounded estimates of the total diet-aging relationship.

The targeted-learning analyses sharpen rather than simply amplify the main finding. The average effects estimated with targeted maximum likelihood estimation and overlap-targeted augmented inverse-probability weighting (AIPW) were directionally consistent with the conventional regression estimates. These analyses are useful because they allow flexible modeling of the exposure and outcome mechanisms and provide a doubly robust estimand under the stated identification assumptions.^31^ They do not, however, make an observational exposure equivalent to randomization. Residual confounding remains possible, particularly for health consciousness, access to healthy food, health-care engagement and unmeasured dimensions of socioeconomic advantage.

Equally important, flexible causal forests did not identify stable heterogeneity. The primary held-out RATE/AUTOC analyses were null across all three aging measures, top-versus-bottom predicted-benefit comparisons were null after multiplicity correction, and repeated random splits failed to reproduce a consistent subgroup signal. This negative result is informative. Precision nutrition is often motivated by the premise that average effects obscure large interindividual differences, but flexible models can generate apparently distinct benefit profiles even when the ranking is not transportable beyond the data used to construct it. Our results provide stronger evidence for a population-average association than for a reliably identifiable subgroup in whom MIND concordance has a substantially different relationship with epigenetic aging. Larger samples and independent validation would be required before any treatment-effect ranking could support individualized dietary recommendations.

The study has several strengths. Diet was measured before DNA methylation, improving temporal ordering relative to existing cross-sectional diet-clock studies. HRS provides a socioeconomically and racially diverse population-based US aging cohort with detailed survey, nutrition and biomarker data.^13^ We analyzed complementary aging measures, incorporated wealth and smoking history into the primary confounder set, separated adiposity adjustment from the primary model, addressed measured selection into the methylation subsample and corrected the primary clock analyses for multiple testing. The exploratory heterogeneous-effect analysis also separated treatment-effect learning from held-out evaluation and examined stability across additional random splits rather than interpreting a single causal-forest ranking at face value.

Several limitations warrant emphasis. First, because the study is observational, the findings cannot establish a causal effect of MIND diet concordance on epigenetic aging. Nonetheless, the observed associations provide important evidence regarding potential biological pathways that warrant further investigation in longer duration intervention studies.^10^ Second, diet was measured once using a self-reported food-frequency questionnaire and may not represent long-term dietary trajectories. However, longitudinal studies of older adult populations have generally reported substantial stability in dietary quality or broad concordance with a dietary pattern over three to 10 years.^32–36^ Third, DNA methylation was measured at a single later assessment, so the outcomes represent subsequent levels of epigenetic aging rather than within-person change from before to after dietary assessment. Fourth, participation in both the nutrition supplement and venous-blood study was selective; inverse-probability weighting can address only measured predictors of selection. Fifth, we used HRS-derived clock measures rather than raw methylation data and did not directly model leukocyte composition or technical batch factors in the outcome regressions. Sixth, alcohol has been linked independently to epigenetic age acceleration,^37^ and because the MIND score includes a wine component, its contribution may be difficult to distinguish from that of overall MIND concordance in this analysis. Finally, the linked HRS clock product available for this analysis contained the original GrimAge and DunedinPoAm38 measures rather than the newer GrimAge2 and DunedinPACE supplement.^20^ Replication with newer clocks, repeated methylation measures and independent cohorts will be important.

Greater MIND diet concordance was therefore associated with lower subsequent epigenetic aging across three complementary methylation measures in middle-aged and older US adults. The convergence of conventional, selection-weighted, and doubly robust estimates supports a consistent average association, whereas the absence of reproducible heterogeneity offered no basis for subgroup-specific targeting. These findings position biological aging as an intermediate phenotype connecting diet quality with healthy aging. Molecular-aging measures merit evaluation as candidate shorter-term endpoints in dietary intervention studies, with attention to their responsiveness to dietary change and their cost relative to clinical outcomes.

## Online Methods

### Study design and data sources

The HRS is a nationally representative, biennial longitudinal study of US adults older than 50 years and their spouses or partners, using a multistage area-probability design with oversampling of Black and Hispanic adults.^38^ The present analysis linked data from the 2012 HRS core interview, the 2013 Health Care and Nutrition Study (HCNS) and DNA methylation-derived epigenetic clocks from the 2016 Venous Blood Study.

The HCNS was fielded from late 2013 through early 2014 and collected dietary information using a self-administered semiquantitative food-frequency questionnaire based on the Harvard food-frequency questionnaire.^28,29^ DNA methylation was assayed from venous blood collected during the 2016 HRS Venous Blood Study, from which HRS constructed a panel of epigenetic clocks.^39^

Among 7,608 HCNS respondents with a derived MIND score and total energy intake, 1,754 matched to the epigenetic-clock file before dietary energy exclusions. We excluded observations with implausible total energy intake using prespecified sex-specific thresholds of <500 or >3,600 kcal daily for women and <600 or >4,200 kcal daily for men. These exclusions were applied in the HCNS source cohort before restriction to DNA methylation participants. The resulting descriptive DNA methylation sample included 1,740 participants.

### MIND diet assessment

MIND concordance was quantified using the established 15-component score.^4,5^ MIND concordance was quantified using the established 15-component score.4,5 Ten components reflect brain-healthy food groups: green leafy vegetables, other vegetables, berries, nuts, whole grains, fish, beans, poultry, olive oil, and wine. Higher intake earns a higher score for nine of these, whereas the wine component assigns its highest score to moderate consumption of about one glass per day. Five components reflect less healthy food groups for which lower intake earns a higher score: red meat, butter or margarine, cheese, pastries and sweets, and fried or fast food. Component scores were summed so that higher values indicate greater concordance.

The primary exposure was the continuous MIND score standardized within the DNA methylation analytic sample. One s.d. corresponded to approximately 1.9 MIND points. Sample-based tertiles were used for secondary analyses and graphical presentation, with the lowest tertile as the reference group.

### Epigenetic-aging measures

We examined three DNA methylation-based aging measures available in the HRS 2016 epigenetic-clock product. PhenoAge was developed to capture methylation patterns associated with a phenotypic-age index derived from clinical biomarkers and mortality.^19^ GrimAge incorporates DNA methylation surrogates of smoking pack-years and mortality-associated plasma proteins.^25^ For PhenoAge and GrimAge, age acceleration was calculated as the residual from a linear regression of DNA methylation age on chronological age at the 2016 blood assessment. Positive residuals represent older epigenetic age than expected for chronological age and negative residuals represent younger epigenetic age than expected.

The third outcome was methylation Pace of Aging, corresponding to the original DunedinPoAm38 algorithm in the HRS clock file.^21^ DunedinPoAm38 is a rate-like measure scaled around approximately 1.0, with higher values indicating a faster estimated pace of physiological aging.

### Covariates

Covariates were selected given their relationships with dietary behavior and epigenetic aging and were measured before or close to the 2013 dietary assessment. From the 2012 HRS core interview, we included age; sex; race and ethnicity, categorized as non-Hispanic White, non-Hispanic Black, Hispanic or non-Hispanic other; educational attainment, categorized as less than high school/GED, high-school graduate, some college or college and higher; household wealth; marital or partner status; smoking status; physical activity; and body mass index.

Household wealth was categorized as zero or negative wealth and quartiles among participants with positive wealth. Smoking was classified as never, former or currently smoking using lifetime and current-smoking variables. Participants were classified as physically active if they reported vigorous activity at least weekly or moderate activity more than once per week, consistent with prior HRS epigenetic-aging analyses.^27^ Total energy intake was obtained from the 2013 HCNS. Alcohol consumption was not separately adjusted for because wine is an explicit component of the MIND score.

Body mass index was not included in the primary confounding-control model because long-term diet can influence adiposity and adjustment could remove part of the pathway through which diet is related to biological aging. It was added in an extended model.

### Primary statistical analysis

Participant characteristics were summarized overall and across MIND tertiles using means and s.d. values for continuous variables and numbers and percentages for categorical variables.

For each epigenetic-aging outcome, we fit linear regression models with heteroskedasticity-robust standard errors. Model 1 adjusted for age, sex, and race and ethnicity. Model 2 was designated the primary model and additionally adjusted for educational attainment, household wealth, marital or partner status, smoking status, physical activity and total energy intake. Model 3 additionally adjusted for body mass index.

The primary estimand was the adjusted mean difference in each epigenetic-aging outcome per 1-s.d. higher MIND score. We applied the Benjamini–Hochberg procedure across the three prespecified primary epigenetic-aging outcomes. Secondary analyses modeled MIND tertiles using the Model 2 covariate set and estimated adjusted outcome means for graphical presentation.

Effect modification by sex and age was evaluated by adding MIND-by-sex and MIND-by-age product terms to the primary model. Interaction analyses were prespecified but considered secondary.

### Selection-weighted sensitivity analysis

To address measured selection into the later DNA methylation sample, we modeled the probability of DNA methylation inclusion among participants in the HCNS source cohort. The logistic selection model included MIND score, age, sex, race and ethnicity, education, household wealth, marital status, smoking, physical activity, total energy intake and body mass index and used the HCNS base weight. Among participants with DNA methylation data, we multiplied the HCNS base weight by the inverse predicted probability of DNA methylation inclusion and normalized the resulting weight to a mean of 1. Weighted robust linear regression then repeated the primary Model 2 analysis. This strategy follows the broader principle used in prior HRS nutrition analyses of accounting for selection from the off-year nutrition sample into subsequent longitudinal analytic cohorts.^40^

### Targeted-learning and heterogeneous-effect analyses

Exploratory analyses evaluated the contrast between high MIND concordance, defined as the highest sample tertile, and lower concordance, defined as the lower two tertiles. A sensitivity contrast compared the highest with the lowest tertile only. The primary adjustment set included age, sex, race and ethnicity, education, household wealth, marital status, smoking status, physical activity and total energy intake. Body mass index was added only in an extended average-effect sensitivity analysis.

We fit honest causal forests using the generalized random forest framework.^41^ Average contrasts were estimated using targeted maximum likelihood estimation with generalized-random-forest nuisance estimates; an overlap-targeted augmented inverse-probability-weighted (AIPW) estimate was calculated as a positivity sensitivity analysis.^31,41^ Outcomes were internally sign-reversed for the forest analyses so that positive predicted benefit corresponded to a lower epigenetic-aging measure under high MIND concordance; reported average estimates were converted back to the original outcome scale.

To evaluate heterogeneity without assessing a prioritization rule in the same observations used to construct it, the sample was split 50:50 within treatment groups into training and evaluation sets. A causal forest in the training sample generated a treatment-benefit ranking for participants in the held-out sample, and a separate causal forest was fit in the evaluation sample. Heterogeneity was quantified using the rank-weighted average treatment effect and the area under the targeting operator characteristic curve (AUTOC), with 500 bootstrap replications.^42^ We also estimated effects in the top and bottom 20% of predicted benefit and compared doubly robust scores between these groups.

To assess sensitivity to a single random split, we repeated the held-out prioritization analysis across five additional stratified 50:50 random splits, using 1,500 trees per forest and 250 bootstrap replications per RATE analysis. We summarized the median and interquartile range of AUTOC estimates, the proportion of positive AUTOC estimates and the proportion of nominal P values below 0.05. Benjamini–Hochberg false-discovery-rate adjustment was applied across the exploratory heterogeneity tests. Causal-forest variable importance was treated as descriptive and was not used to define subgroups post hoc.

These targeted-learning analyses estimate causal contrasts only under the assumptions of consistency, conditional exchangeability given the measured covariates and positivity. Because concordance with the MIND diet was observational rather than randomized, we treat these analyses as exploratory and do not interpret them as equivalent to trial effects.

### Missing data and software

Analyses used complete cases for variables included in each model. The primary regression models included 1,722 participants. The body-mass-index-extended models included 1,702 participants. The main targeted-learning contrast included 1,722 participants, and the highest-versus-lowest-tertile sensitivity contrast included 1,226 participants.

Primary analyses were conducted in Stata version 18.5 (StataCorp). Targeted-learning and generalized-random-forest analyses were conducted in R version 4.6.1 using grf version 2.6.1. Stata extended missing-value codes were explicitly treated as missing on transfer to R.

## Ethics

HRS procedures are reviewed and approved by the University of Michigan institutional review board. Participants provide informed consent for study participation, including written consent for venous blood collection. This secondary analysis used HRS data under applicable data use agreements.

## Data availability

The Health and Retirement Study included in this study can be accessed via its website (https://hrs.isr.umich.edu/data-products). DNA methylation and epigenetic-clock data are sensitive HRS data products and require approved access and applicable data use agreements. The derived analytic variables used in this study can be recreated from the HRS products described above.

## Code availability

All Stata and R scripts used to construct the analytic cohort, reproduce the statistical analyses and generate source data for the figures will be available upon request.

## Acknowledgements

The HRS is sponsored by the National Institute on Aging (U01AG009740) and the Social Security Administration and is conducted by the University of Michigan.

## Author contributions

J.W., V.H.C.W., and D.Z. contributed to the study design, analytic strategy, and drafted the manuscript. J.W. and V.H.C.W. contributed to data cleaning. J.W. conducted the statistical analysis and performed supplementary analyses. V.H.C.W. supported data visualization. All authors contributed to the interpretation of results and helped critically revise the manuscript.

## Additional information

A Nature Portfolio Reporting Summary will be provided with the submission.

## Extended Data and Supplementary Information

## Extended Data and Supplementary Information

**Extended Data Table 1.** Participant characteristics by MIND concordance tertile. Includes age, sex, race and ethnicity, education, household wealth, marital or partner status, smoking status, physical activity, body mass index, energy intake and MIND score.

|  | MIND diet concordance |  |  |  |
|---|---|---|---|---|
|  | Low | Middle | High | Total |
| N | 748 (43.0%) | 501 (28.8%) | 491 (28.2%) | 1,740 (100.0%) |
| Age, y | 66.9 (9.3) | 65.9 (9.7) | 64.7 (9.5) | 66.0 (9.5) |
| Sex |  |  |  |  |
| Male | 359 (48.0%) | 198 (39.5%) | 150 (30.5%) | 707 (40.6%) |
| Female | 389 (52.0%) | 303 (60.5%) | 341 (69.5%) | 1,033 (59.4%) |
| Race/ethnicity |  |  |  |  |
| Non-Hispanic White | 555 (74.2%) | 337 (67.3%) | 339 (69.2%) | 1,231 (70.8%) |
| Non-Hispanic Black | 118 (15.8%) | 78 (15.6%) | 56 (11.4%) | 252 (14.5%) |
| Hispanic | 58 (7.8%) | 68 (13.6%) | 80 (16.3%) | 206 (11.8%) |
| Non-Hispanic Other | 17 (2.3%) | 18 (3.6%) | 15 (3.1%) | 50 (2.9%) |
| Educational attainment |  |  |  |  |
| <HS/GED | 164 (22.0%) | 93 (18.6%) | 79 (16.1%) | 336 (19.3%) |
| HS graduate | 270 (36.1%) | 145 (28.9%) | 99 (20.2%) | 514 (29.6%) |
| Some college | 200 (26.8%) | 127 (25.3%) | 119 (24.2%) | 446 (25.6%) |
| College+ | 113 (15.1%) | 136 (27.1%) | 194 (39.5%) | 443 (25.5%) |
| Household wealth |  |  |  |  |
| Zero/negative | 89 (11.9%) | 51 (10.2%) | 44 (9.0%) | 184 (10.6%) |
| Q1 | 185 (24.8%) | 103 (20.7%) | 88 (18.0%) | 376 (21.7%) |
| Q2 | 199 (26.7%) | 105 (21.1%) | 93 (19.0%) | 397 (22.9%) |
| Q3 | 156 (20.9%) | 107 (21.5%) | 120 (24.5%) | 383 (22.1%) |
| Q4 | 116 (15.6%) | 132 (26.5%) | 145 (29.6%) | 393 (22.7%) |
| Marital status |  |  |  |  |
| Not married | 264 (35.4%) | 163 (32.8%) | 174 (35.5%) | 601 (34.7%) |
| Married/partnered | 481 (64.6%) | 334 (67.2%) | 316 (64.5%) | 1,131 (65.3%) |
| Smoking status |  |  |  |  |
| Never | 306 (41.4%) | 235 (47.3%) | 236 (48.2%) | 777 (45.0%) |
| Former | 309 (41.8%) | 214 (43.1%) | 234 (47.8%) | 757 (43.9%) |
| Current | 124 (16.8%) | 48 (9.7%) | 20 (4.1%) | 192 (11.1%) |
| Physically active |  |  |  |  |
| Inactive | 353 (47.4%) | 166 (33.3%) | 103 (21.0%) | 622 (35.9%) |
| Active | 391 (52.6%) | 332 (66.7%) | 387 (79.0%) | 1,110 (64.1%) |
| Body mass index, kg/m<sup>2</sup> | 29.5 (6.0) | 29.0 (6.0) | 27.9 (5.7) | 28.9 (5.9) |
| Energy intake, kcal/d | 1744.7 (692.9) | 1723.0 (686.3) | 1873.1 (633.2) | 1774.7 (677.1) |
| MIND diet score | 5.4 (1.0) | 7.5 (0.4) | 9.5 (1.0) | 7.2 (1.9) |

Values are mean (SD) or No. (%). No hypothesis tests are shown for descriptive characteristics.

**Extended Data Table 2.**
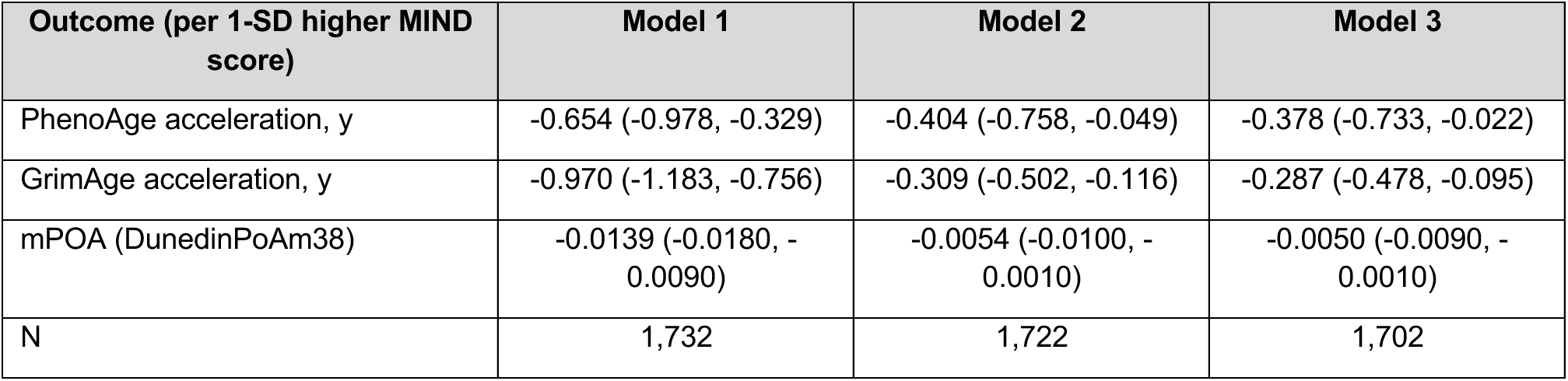
Adjusted associations of continuous MIND score with epigenetic aging. *Model 1: age, sex and race and ethnicity. Model 2: Model 1 plus education, household wealth, marital or partner status, smoking status, physical activity and energy intake. Model 3: Model 2 plus body mass index*.

| **Outcome (per 1-SD higher MIND score)** | **Model 1** | **Model 2** | **Model 3** |
|---|---|---|---|
| PhenoAge acceleration, y | -0.654 (-0.978, -0.329) | -0.404 (-0.758, -0.049) | -0.378 (-0.733, -0.022) |
| GrimAge acceleration, y | -0.970 (-1.183, -0.756) | -0.309 (-0.502, -0.116) | -0.287 (-0.478, -0.095) |
| mPOA (DunedinPoAm38) | -0.0139 (-0.0180, -0.0090) | -0.0054 (-0.0100, -0.0010) | -0.0050 (-0.0090, -0.0010) |
| N | 1,732 | 1,722 | 1,702 |

**Extended Data Table 3.** Selection-weighted sensitivity analyses. *Associations of continuous MIND score with epigenetic aging after weighting for measured selection from the 2013 nutrition cohort into the 2016 DNA methylation subsample. The selection model incorporated MIND score, age, sex, race and ethnicity, education, household wealth, marital status, smoking, physical activity, energy intake and body mass index, and was combined with the Health Care and Nutrition Study base weight*.

| **Outcome (per 1-SD higher MIND score)** | **β (selection-weighted)** | **95% CI** | **N** |
|---|---|---|---|
| PhenoAge acceleration, y | -0.714 | -1.177, -0.250 | 1,644 |
| GrimAge acceleration, y | -0.376 | -0.604, -0.148 | 1,644 |
| mPOA (DunedinPoAm38) | -0.0082 | -0.0130, -0.0030 | 1,644 |

**Extended Data Table 4.** Targeted maximum likelihood, overlap-AIPW and body-mass-index-adjusted average estimates. *Exploratory targeted-learning estimates of the average association between high MIND concordance and epigenetic aging. Negative original-scale values indicate lower epigenetic aging with high MIND concordance. Panel a shows the primary contrast (highest MIND tertile vs. lower two tertiles); panel b shows the sensitivity contrast restricted to the highest vs. lowest tertile*.

| Outcome | Estimator | N (group / group) | Estimate (95% CI) | P (FDR q) |
|---|---|---|---|---|
| **Panel: a — Highest tertile vs. lower two tertiles (primary contrast)** |  |  |  |  |
| PhenoAge acceleration, y | TMLE (primary covariates) | 1722 (489 high / 1233 lower) | -0.894 (-1.663, -0.126) | 0.023 (0.023) |
| PhenoAge acceleration, y | Overlap-AIPW (primary covariates) | 1722 (489 high / 1233 lower) | -0.817 (-1.543, -0.092) | 0.027 (0.027) |
| PhenoAge acceleration, y | TMLE (+ body mass index) | 1702 (485 high / 1217 lower) | -0.894 (-1.655, -0.133) | 0.021 (0.021) |
| GrimAge acceleration, y | TMLE (primary covariates) | 1722 (489 high / 1233 lower) | -0.847 (-1.268, -0.426) | <0.001 (<0.001) |
| GrimAge acceleration, y | Overlap-AIPW (primary covariates) | 1722 (489 high / 1233 lower) | -0.766 (-1.149, -0.382) | <0.001 (<0.001) |
| GrimAge acceleration, y | TMLE (+ body mass index) | 1702 (485 high / 1217 lower) | -0.782 (-1.197, -0.367) | <0.001 (0.001) |
| mPOA (DunedinPoAm38) | TMLE (primary covariates) | 1722 (489 high / 1233 lower) | -0.0125 (-0.0223, -0.0027) | 0.012 (0.015) |
| mPOA (DunedinPoAm38) | Overlap-AIPW (primary covariates) | 1722 (489 high / 1233 lower) | -0.0114 (-0.0205, -0.0024) | 0.013 (0.016) |
| mPOA (DunedinPoAm38) | TMLE (+ body mass index) | 1702 (485 high / 1217 lower) | -0.0115 (-0.0212, -0.0018) | 0.020 (0.021) |
| **Panel: b — Highest tertile vs. lowest tertile (sensitivity contrast)** |  |  |  |  |
| **Outcome** | **Estimator** | **N (group / group)** | **Estimate (95% CI)** | **P (FDR q)** |
| PhenoAge acceleration, y | TMLE (primary covariates) | 1226 (489 high / 737 lower) | -1.119 (-1.984, -0.255) | 0.011 (0.015) |
| PhenoAge acceleration, y | Overlap-AIPW (primary covariates) | 1226 (489 high / 737 lower) | -1.101 (-1.927, -0.275) | 0.009 (0.013) |
| PhenoAge acceleration, y | TMLE (+ body mass index) | 1219 (485 high / 734 lower) | -1.047 (-1.901, -0.192) | 0.016 (0.021) |
| GrimAge acceleration, y | TMLE (primary covariates) | 1226 (489 high / 737 lower) | -0.866 (-1.351, -0.382) | <0.001 (0.001) |
| GrimAge acceleration, y | Overlap-AIPW (primary covariates) | 1226 (489 high / 737 lower) | -0.795 (-1.244, -0.345) | <0.001 (0.002) |
| GrimAge acceleration, y | TMLE (+ body mass index) | 1219 (485 high / 734 lower) | -0.785 (-1.259, -0.312) | 0.001 (0.003) |
| mPOA (DunedinPoAm38) | TMLE (primary covariates) | 1226 (489 high / 737 lower) | -0.0156 (-0.0267, -0.0045) | 0.006 (0.012) |
| mPOA (DunedinPoAm38) | Overlap-AIPW (primary covariates) | 1226 (489 high / 737 lower) | -0.0147 (-0.0251, -0.0043) | 0.006 (0.011) |
| mPOA (DunedinPoAm38) | TMLE (+ body mass index) | 1219 (485 high / 734 lower) | -0.0148 (-0.0256, -0.0039) | 0.008 (0.015) |

**Extended Data Fig. 1.**
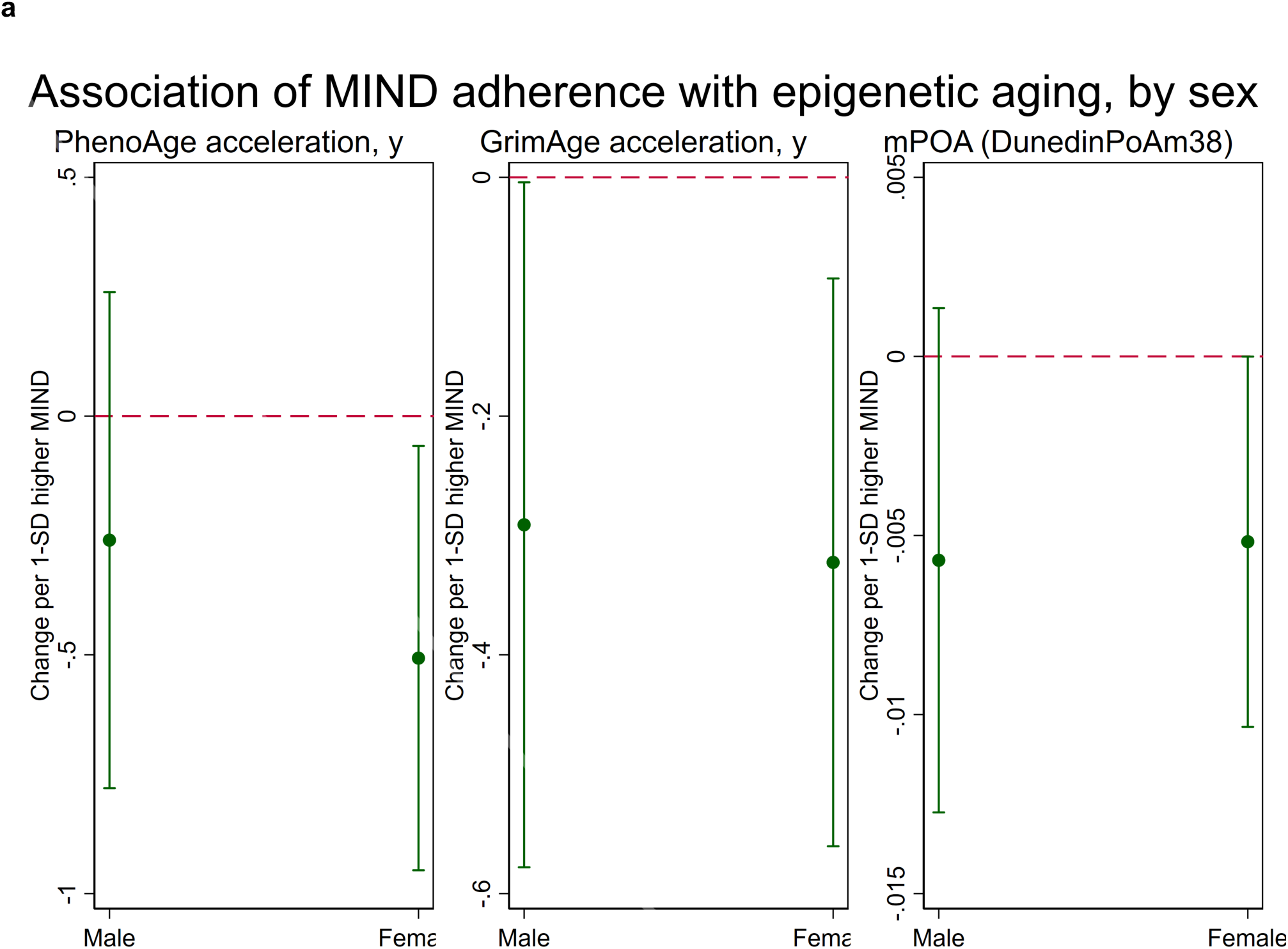
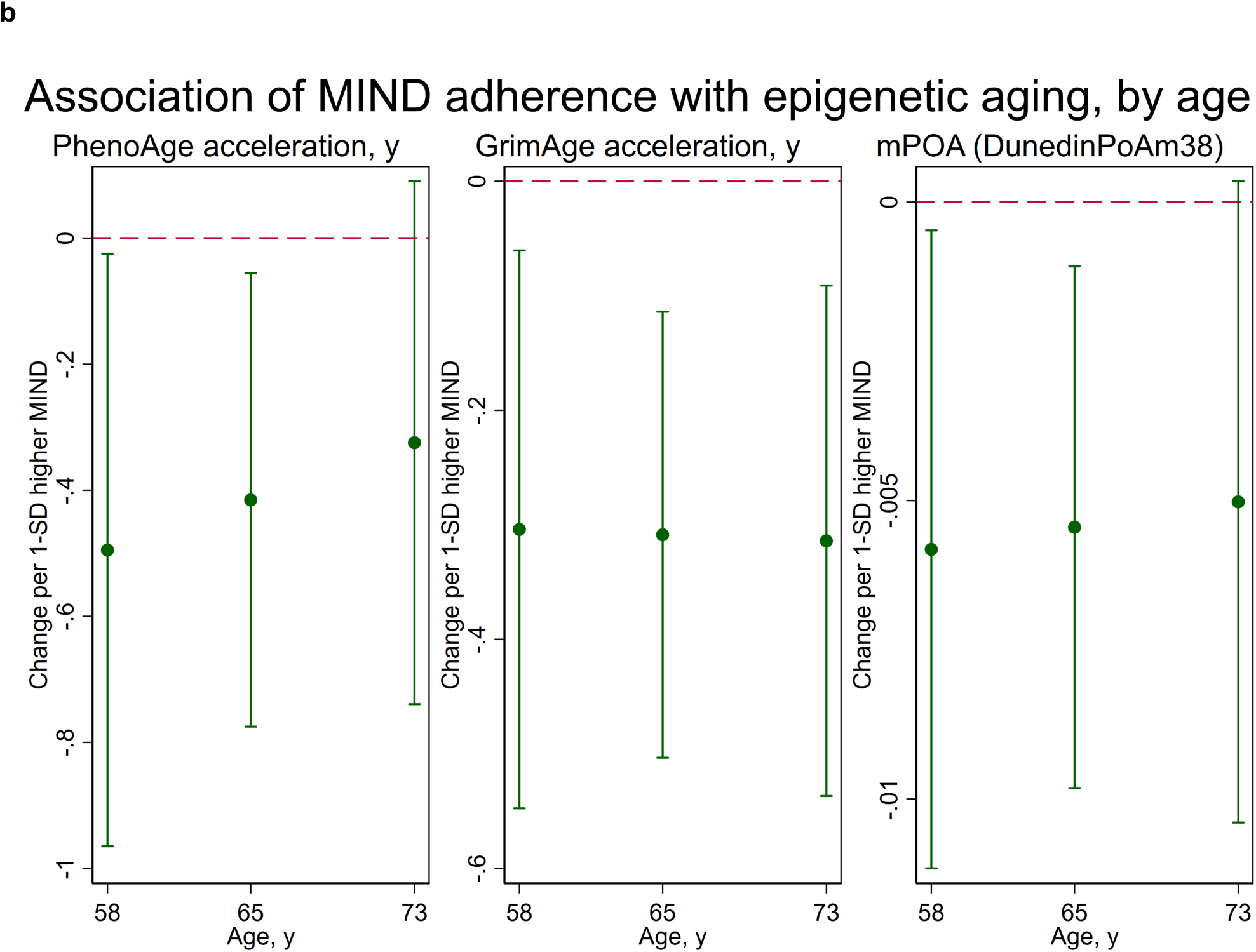
Associations of MIND concordance with epigenetic aging by sex and age. Adjusted mean differences (points) and 95% confidence intervals (error bars) in PhenoAge acceleration, GrimAge acceleration and methylation Pace of Aging per 1-s.d. higher MIND score. a, Estimates stratified by sex (male, female). b, Estimates stratified by age (58, 65 and 73 years, representing approximately the 16th, 50th and 84th percentiles of the age distribution). Corresponding interaction P values for sex and age are reported in Supplementary Table 1.

**Supplementary Table 1.** Interaction P values for age and sex. P values for MIND-score-by-sex and MIND-score-by-age interaction terms, added individually to the primary multivariable model for each epigenetic-aging outcome.

| **Outcome** | **P (MIND × sex)** | **P (MIND × age)** |
|---|---|---|
| PhenoAge acceleration, y | 0.454 | 0.525 |
| GrimAge acceleration, y | 0.860 | 0.940 |
| mPOA (DunedinPoAm38) | 0.904 | 0.802 |

**Supplementary Table 2.**
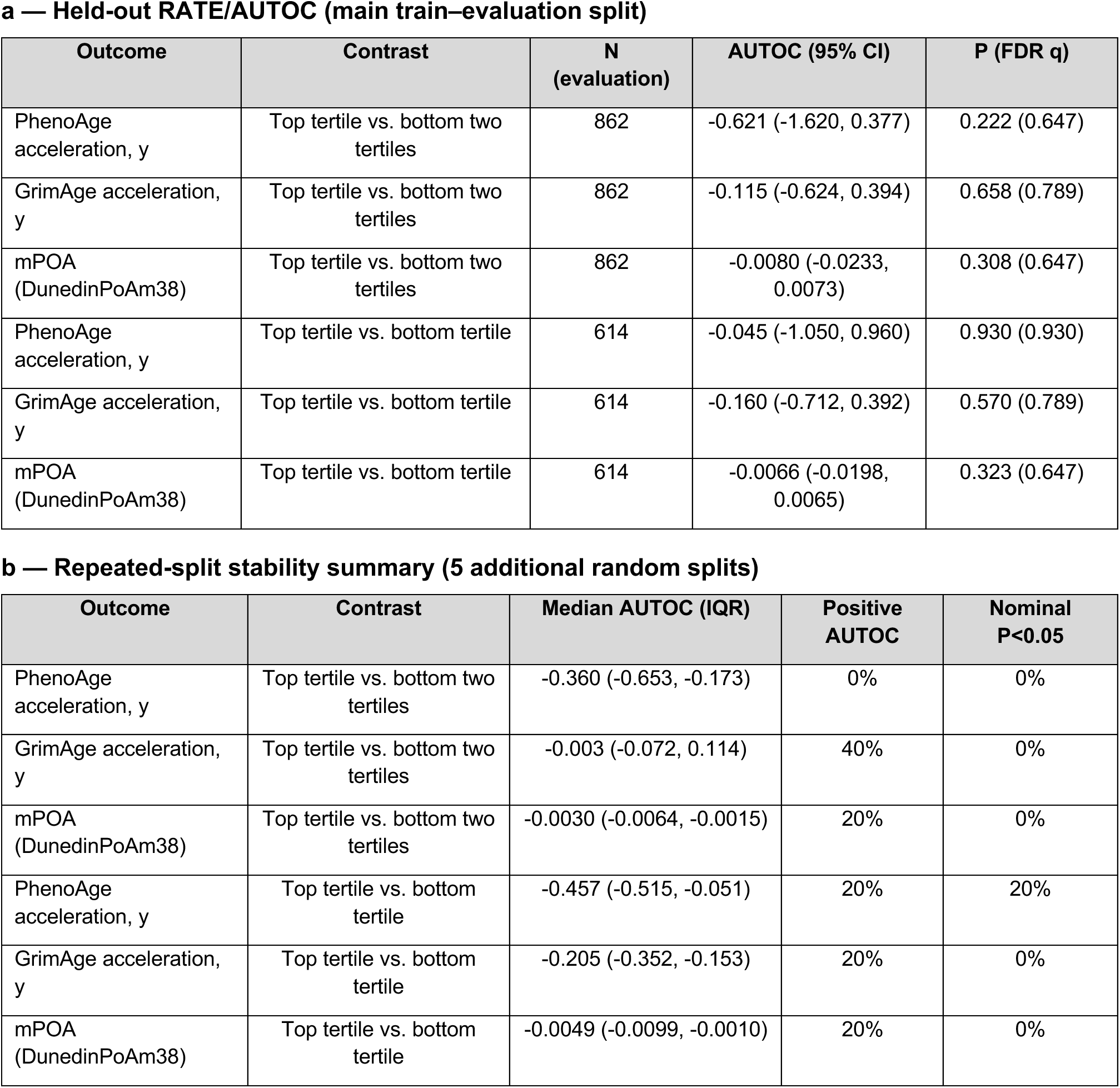
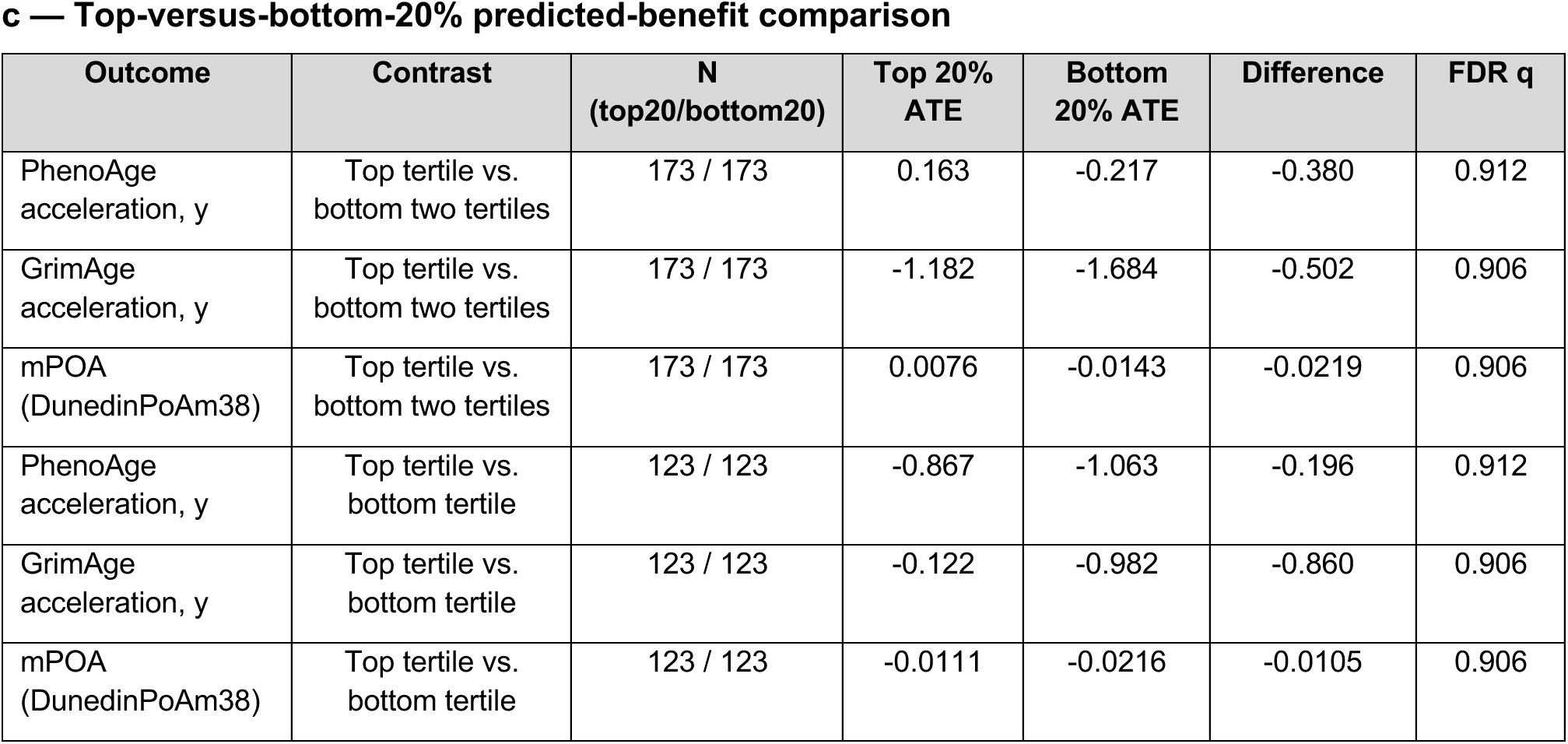
Held-out RATE/AUTOC, repeated-split stability and top-versus-bottom-20% heterogeneous-effect analyses. Causal-forest evaluation of treatment-effect heterogeneity for high MIND concordance. Panel a, primary held-out rank-weighted average treatment effect (AUTOC) test. Panel b, stability of the AUTOC test across five additional random train–evaluation splits (median, interquartile range, proportion of positive AUTOC estimates and proportion of nominally significant [P<0.05] splits). Panel c, direct comparison of predicted benefit between the top and bottom 20% of the held-out sample.

| a — Held-out RATE/AUTOC (main train–evaluation split) |  |  |  |  |  |  |
|---|---|---|---|---|---|---|
| Outcome | Contrast | N (evaluation) | AUTOC (95% CI) | P (FDR q) |  |  |
| PhenoAge acceleration, y | Top tertile vs. bottom two tertiles | 862 | -0.621 (-1.620, 0.377) | 0.222 (0.647) |  |  |
| GrimAge acceleration, y | Top tertile vs. bottom two tertiles | 862 | -0.115 (-0.624, 0.394) | 0.658 (0.789) |  |  |
| mPOA (DunedinPoAm38) | Top tertile vs. bottom two tertiles | 862 | -0.0080 (-0.0233, 0.0073) | 0.308 (0.647) |  |  |
| PhenoAge acceleration, y | Top tertile vs. bottom tertile | 614 | -0.045 (-1.050, 0.960) | 0.930 (0.930) |  |  |
| GrimAge acceleration, y | Top tertile vs. bottom tertile | 614 | -0.160 (-0.712, 0.392) | 0.570 (0.789) |  |  |
| mPOA (DunedinPoAm38) | Top tertile vs. bottom tertile | 614 | -0.0066 (-0.0198, 0.0065) | 0.323 (0.647) |  |  |
| b — Repeated-split stability summary (5 additional random splits) |  |  |  |  |  |  |
| Outcome | Contrast | Median AUTOC (IQR) | Positive AUTOC | Nominal P<0.05 |  |  |
| PhenoAge acceleration, y | Top tertile vs. bottom two tertiles | -0.360 (-0.653, -0.173) | 0% | 0% |  |  |
| GrimAge acceleration, y | Top tertile vs. bottom two tertiles | -0.003 (-0.072, 0.114) | 40% | 0% |  |  |
| mPOA (DunedinPoAm38) | Top tertile vs. bottom two tertiles | -0.0030 (-0.0064, -0.0015) | 20% | 0% |  |  |
| PhenoAge acceleration, y | Top tertile vs. bottom tertile | -0.457 (-0.515, -0.051) | 20% | 20% |  |  |
| GrimAge acceleration, y | Top tertile vs. bottom tertile | -0.205 (-0.352, -0.153) | 20% | 0% |  |  |
| mPOA (DunedinPoAm38) | Top tertile vs. bottom tertile | -0.0049 (-0.0099, -0.0010) | 20% | 0% |  |  |
| c — Top-versus-bottom-20% predicted-benefit comparison |  |  |  |  |  |  |
| Outcome | Contrast | N (top20/bottom20) | Top 20% ATE | Bottom 20% ATE | Difference | FDR q |
| PhenoAge acceleration, y | Top tertile vs. bottom two tertiles | 173 / 173 | 0.163 | -0.217 | -0.380 | 0.912 |
| GrimAge acceleration, y | Top tertile vs. bottom two tertiles | 173 / 173 | -1.182 | -1.684 | -0.502 | 0.906 |
| mPOA (DunedinPoAm38) | Top tertile vs. bottom two tertiles | 173 / 173 | 0.0076 | -0.0143 | -0.0219 | 0.906 |
| PhenoAge acceleration, y | Top tertile vs. bottom tertile | 123 / 123 | -0.867 | -1.063 | -0.196 | 0.912 |
| GrimAge acceleration, y | Top tertile vs. bottom tertile | 123 / 123 | -0.122 | -0.982 | -0.860 | 0.906 |
| mPOA (DunedinPoAm38) | Top tertile vs. bottom tertile | 123 / 123 | -0.0111 | -0.0216 | -0.0105 | 0.906 |

**Supplementary Table 3.** Causal-forest variable importance and participant profiles across predicted-benefit groups. Panel a, causal-forest variable importance (descriptive; not used for inference) for the primary contrast (highest MIND tertile vs. lower two tertiles), ranked within outcome. Panel b, characteristics of participants in the top and bottom 20% of predicted benefit for the same contrast. Standardized mean differences (SMD) are shown for the continuous variables age and energy intake; differences in percentage points are shown for categorical variables.

| a — Causal-forest variable importance (highest tertile vs. lower two tertiles) |  |  |  |  |
|---|---|---|---|---|
| PhenoAge acceleration, y |  |  |  |  |
| **Rank** | **Variable** | **Importance** |  |  |
| 1 | Total energy intake | 0.419 |  |  |
| 2 | Age | 0.258 |  |  |
| 3 | Household wealth category (level 3) | 0.042 |  |  |
| 4 | Married or partnered | 0.034 |  |  |
| 5 | Education category (level 4) | 0.030 |  |  |
| 6 | Household wealth category (level 4) | 0.029 |  |  |
| 7 | Smoking status (level 1) | 0.029 |  |  |
| 8 | Education category (level 3) | 0.029 |  |  |
| 9 | Race and ethnicity (level 1) | 0.025 |  |  |
| 10 | Physically active | 0.023 |  |  |
| 11 | Household wealth category (level 2) | 0.020 |  |  |
| 12 | Sex: female | 0.017 |  |  |
| 13 | Race and ethnicity (level 2) | 0.016 |  |  |
| 14 | Household wealth category (level 1) | 0.014 |  |  |
| 15 | Education category (level 2) | 0.012 |  |  |
| 16 | Smoking status (level 2) | 0.002 |  |  |
| 17 | Race and ethnicity (level 3) | 0.000 |  |  |
| GrimAge acceleration, y |  |  |  |  |
| **Rank** | **Variable** | **Importance** |  |  |
| 1 | Total energy intake | 0.302 |  |  |
| 2 | Age | 0.273 |  |  |
| 3 | Education category (level 3) | 0.096 |  |  |
| 4 | Education category (level 4) | 0.079 |  |  |
| 5 | Race and ethnicity (level 1) | 0.042 |  |  |
| 6 | Household wealth category (level 4) | 0.035 |  |  |
| 7 | Household wealth category (level 2) | 0.029 |  |  |
| 8 | Household wealth category (level 3) | 0.024 |  |  |
| 9 | Married or partnered | 0.022 |  |  |
| 10 | Smoking status (level 1) | 0.021 |  |  |
| 11 | Household wealth category (level 1) | 0.018 |  |  |
| 12 | Sex: female | 0.017 |  |  |
| 13 | Race and ethnicity (level 2) | 0.016 |  |  |
| 14 | Physically active | 0.015 |  |  |
| 15 | Education category (level 2) | 0.008 |  |  |
| 16 | Smoking status (level 2) | 0.004 |  |  |
| 17 | Race and ethnicity (level 3) | 0.000 |  |  |
| mPOA (DunedinPoAm38) |  |  |  |  |
| **Rank** | **Variable** | **Importance** |  |  |
| 1 | Total energy intake | 0.309 |  |  |
| 2 | Age | 0.303 |  |  |
| 3 | Race and ethnicity (level 2) | 0.059 |  |  |
| 4 | Education category (level 4) | 0.058 |  |  |
| 5 | Education category (level 3) | 0.042 |  |  |
| 6 | Physically active | 0.031 |  |  |
| 7 | Smoking status (level 1) | 0.030 |  |  |
| 8 | Household wealth category (level 2) | 0.030 |  |  |
| 9 | Household wealth category (level 4) | 0.030 |  |  |
| 10 | Race and ethnicity (level 1) | 0.026 |  |  |
| 11 | Household wealth category (level 3) | 0.022 |  |  |
| 12 | Married or partnered | 0.018 |  |  |
| 13 | Sex: female | 0.016 |  |  |
| 14 | Education category (level 2) | 0.014 |  |  |
| 15 | Household wealth category (level 1) | 0.010 |  |  |
| 16 | Smoking status (level 2) | 0.003 |  |  |
| 17 | Race and ethnicity (level 3) | 0.000 |  |  |
| b — Participant profiles, top vs. bottom 20% predicted benefit (highest tertile vs. lower two tertiles) |  |  |  |  |
| PhenoAge acceleration, y |  |  |  |  |
| **Variable** | **Level** | **Top 20%** | **Bottom 20%** | **Difference** |
| Age, y | — | 67.75 (7.86) | 63.11 (9.52) | SMD = 0.531 |
| Total energy intake | — | 1084.20 (222.17) | 2078.80 (334.80) | SMD = -3.501 |
| Sex | 0 | 32.9% | 46.8% | -13.9 pp |
| Sex | 1 | 67.1% | 53.2% | 13.9 pp |
| Race and ethnicity | 0 | 69.4% | 69.9% | -0.6 pp |
| Race and ethnicity | 1 | 15.0% | 17.3% | -2.3 pp |
| Race and ethnicity | 2 | 9.8% | 10.4% | -0.6 pp |
| Race and ethnicity | 3 | 5.8% | 2.3% | 3.5 pp |
| Education category | 1 | 23.7% | 13.9% | 9.8 pp |
| Education category | 2 | 30.6% | 32.9% | -2.3 pp |
| Education category | 3 | 20.2% | 26.6% | -6.4 pp |
| Education category | 4 | 25.4% | 26.6% | -1.2 pp |
| Household wealth category | 0 | 8.7% | 10.4% | -1.7 pp |
| Household wealth category | 1 | 23.7% | 17.3% | 6.4 pp |
| Household wealth category | 2 | 33.5% | 27.2% | 6.4 pp |
| Household wealth category | 3 | 1.7% | 41.0% | -39.3 pp |
| Household wealth category | 4 | 32.4% | 4.0% | 28.3 pp |
| Married or partnered | 0 | 33.5% | 26.6% | 6.9 pp |
| Married or partnered | 1 | 66.5% | 73.4% | -6.9 pp |
| Smoking status | 0 | 54.3% | 31.2% | 23.1 pp |
| Smoking status | 1 | 38.7% | 58.4% | -19.7 pp |
| Smoking status | 2 | 6.9% | 10.4% | -3.5 pp |
| Physically active | 0 | 48.6% | 35.8% | 12.7 pp |
| Physically active | 1 | 51.4% | 64.2% | -12.7 pp |
| GrimAge acceleration, y |  |  |  |  |
| **Variable** | **Level** | **Top 20%** | **Bottom 20%** | **Difference** |
| Age, y | — | 65.45 (10.25) | 61.75 (7.38) | SMD = 0.415 |
| Total energy intake | — | 1403.38 (480.94) | 2210.34 (579.36) | SMD = -1.516 |
| Sex | 0 | 24.3% | 49.1% | -24.9 pp |
| Sex | 1 | 75.7% | 50.9% | 24.9 pp |
| Race and ethnicity | 0 | 72.3% | 57.2% | 15.0 pp |
| Race and ethnicity | 1 | 23.7% | 6.4% | 17.3 pp |
| Race and ethnicity | 2 | 1.2% | 34.1% | -32.9 pp |
| Race and ethnicity | 3 | 2.9% | 2.3% | 0.6 pp |
| Education category | 1 | 12.1% | 27.2% | -15.0 pp |
| Education category | 2 | 19.1% | 39.3% | -20.2 pp |
| Education category | 3 | 60.7% | 6.4% | 54.3 pp |
| Education category | 4 | 8.1% | 27.2% | -19.1 pp |
| Household wealth category | 0 | 11.6% | 16.2% | -4.6 pp |
| Household wealth category | 1 | 28.3% | 19.1% | 9.2 pp |
| Household wealth category | 2 | 17.3% | 25.4% | -8.1 pp |
| Household wealth category | 3 | 12.1% | 37.0% | -24.9 pp |
| Household wealth category | 4 | 30.6% | 2.3% | 28.3 pp |
| Married or partnered | 0 | 57.8% | 13.3% | 44.5 pp |
| Married or partnered | 1 | 42.2% | 86.7% | -44.5 pp |
| Smoking status | 0 | 57.2% | 37.0% | 20.2 pp |
| Smoking status | 1 | 24.9% | 52.0% | -27.2 pp |
| Smoking status | 2 | 17.9% | 11.0% | 6.9 pp |
| Physically active | 0 | 39.9% | 34.7% | 5.2 pp |
| Physically active | 1 | 60.1% | 65.3% | -5.2 pp |
| mPOA (DunedinPoAm38) |  |  |  |  |
| **Variable** | **Level** | **Top 20%** | **Bottom 20%** | **Difference** |
| Age, y | — | 63.12 (10.22) | 71.32 (6.75) | SMD = -0.947 |
| Total energy intake | — | 2207.20 (617.14) | 1390.32 (343.58) | SMD = 1.636 |
| Sex | 0 | 39.9% | 47.4% | -7.5 pp |
| Sex | 1 | 60.1% | 52.6% | 7.5 pp |
| Race and ethnicity | 0 | 50.3% | 96.0% | -45.7 pp |
| Race and ethnicity | 1 | 30.6% | 0.0% | 30.6 pp |
| Race and ethnicity | 2 | 15.6% | 2.9% | 12.7 pp |
| Race and ethnicity | 3 | 3.5% | 1.2% | 2.3 pp |
| Education category | 1 | 24.9% | 11.6% | 13.3 pp |
| Education category | 2 | 22.0% | 30.1% | -8.1 pp |
| Education category | 3 | 37.0% | 4.6% | 32.4 pp |
| Education category | 4 | 16.2% | 53.8% | -37.6 pp |
| Household wealth category | 0 | 17.9% | 0.6% | 17.3 pp |
| Household wealth category | 1 | 27.7% | 6.4% | 21.4 pp |
| Household wealth category | 2 | 34.1% | 17.9% | 16.2 pp |
| Household wealth category | 3 | 5.8% | 43.4% | -37.6 pp |
| Household wealth category | 4 | 14.5% | 31.8% | -17.3 pp |
| Married or partnered | 0 | 38.2% | 27.2% | 11.0 pp |
| Married or partnered | 1 | 61.8% | 72.8% | -11.0 pp |
| Smoking status | 0 | 48.6% | 48.0% | 0.6 pp |
| Smoking status | 1 | 38.7% | 43.9% | -5.2 pp |
| Smoking status | 2 | 12.7% | 8.1% | 4.6 pp |
| Physically active | 0 | 39.3% | 35.3% | 4.0 pp |
| Physically active | 1 | 60.7% | 64.7% | -4.0 pp |

## References

1. Kennedy, B. K. et al. Geroscience: linking aging to chronic disease. Cell 159, 709–713 (2014).

2. Abbad-Gomez, D. et al. Dietary patterns and accelerated multimorbidity in older adults. Nature Aging 5, 1481–1490 (2025).

3. Tessier, A.-J. et al. Optimal dietary patterns for healthy aging. Nature Medicine 31, 1644–1652 (2025).

4. Morris, M. C. et al. MIND diet associated with reduced incidence of Alzheimer’s disease. Alzheimers Dement 11, 1007–1014 (2015).

5. Morris, M. C. et al. MIND diet slows cognitive decline with aging. Alzheimers Dement 11, 1015–1022 (2015).

6. Morris, M. C. et al. Nutrients and bioactives in green leafy vegetables and cognitive decline. Neurology 90, e214–e222 (2018).

7. Lamport, D. J. & Williams, C. M. Polyphenols and Cognition In Humans: An Overview of Current Evidence from Recent Systematic Reviews and Meta-Analyses. Brain Plast 6, 139–153.

8. Gu, Y. et al. Mediterranean diet and brain structure in a multiethnic elderly cohort. Neurology 85, 1744–1751 (2015).

9. Agarwal, P. et al. Association of Mediterranean-DASH Intervention for Neurodegenerative Delay and Mediterranean Diets With Alzheimer Disease Pathology. Neurology 100, e2259–e2268 (2023).

10. Barnes, L. L. et al. Trial of the MIND Diet for Prevention of Cognitive Decline in Older Persons. New England Journal of Medicine 389, 602–611 (2023).

11. Fei, L. et al. Mediterranean-Dietary Approaches to Stop Hypertension Intervention for Neurodegeneration Delay (MIND) Diet, Circulating Inflammatory Biomarkers, and Cognitive Function: A Prospective Study. J Nutr 155, 3536–3546 (2025).

12. Upadhyay, R. K. Emerging risk biomarkers in cardiovascular diseases and disorders. J Lipids 2015, 971453 (2015).

13. Halder, D., Prajapati, D., Banerjee, T., Biswas, S. & Ghosh, B. Epigenetic modulators as therapeutics for neurodegenerative disorders/Alzheimer’s disease. Methods Enzymol 733, 163–222 (2026).

14. Bordoni, L., Agostinho de Sousa, J., Zhuo, J. & von Meyenn, F. Evaluating the connection between diet quality, EpiNutrient intake and epigenetic age: an observational study. Am J Clin Nutr 120, 1143–1155 (2024).

15. Behl, T. et al. Epigenetic biomarkers in Alzheimer’s disease: Diagnostic and prognostic relevance. Ageing Res Rev 102, 102556 (2024).

16. Rutledge, J., Oh, H. & Wyss-Coray, T. Measuring biological age using omics data. Nat Rev Genet 23, 715–727 (2022).

17. Levine, M. E. et al. An epigenetic biomarker of aging for lifespan and healthspan. Aging (Albany NY) 10, 573–591 (2018).

18. Lu, A. T. et al. DNA methylation GrimAge strongly predicts lifespan and healthspan. Aging (Albany NY) 11, 303–327 (2019).

19. Levine, M. E. et al. An epigenetic biomarker of aging for lifespan and healthspan. Aging (Albany NY) 10, 573–591 (2018).

20. Belsky, D. W. et al. DunedinPACE, a DNA methylation biomarker of the pace of aging. Elife 11, e73420 (2022).

21. Belsky, D. W. et al. Quantification of the pace of biological aging in humans through a blood test, the DunedinPoAm DNA methylation algorithm. eLife 9, e54870 (2020).

22. Fuentealba, M. et al. A blood-based epigenetic clock for intrinsic capacity predicts mortality and is associated with clinical, immunological and lifestyle factors. Nature Aging 5, 1207–1216 (2025).

23. Grande de França, N., et al. The role of dietary patterns on epigenetic and inflammatory aging based on the INSPIRE-T study. Commun Med (Lond) 6, 410 (2026).

24. Tavares, J. F. Associations between diet quality, epigenetic aging and epigenome in two population-based cohorts. Nature Communications 17, 9232 (2026).

25. Lu, A. T. et al. DNA methylation GrimAge strongly predicts lifespan and healthspan. Aging (Albany NY) 11, 303–327 (2019).

26. Schmitz, L. L. et al. The Socioeconomic Gradient in Epigenetic Ageing Clocks: Evidence from the Multi-Ethnic Study of Atherosclerosis and the Health and Retirement Study. Epigenetics 17, 589–611 (2022).

27. Ammous, F., Peterson, M. D., Mitchell, C. & Faul, J. D. Physical Activity Is Associated With Decreased Epigenetic Aging: Findings From the Health and Retirement Study. J Cachexia Sarcopenia Muscle 16, e13873 (2025).

28. Lupoli, R. et al. Effects of Mediterranean Diet on Endothelial Reactivity in Individuals with High Cardiometabolic Risk: A Randomized Controlled Parallel-Group Preliminary Trial. Biomedicines 12, 2595 (2024).

29. Leitão, C. et al. The Effect of Nutrition on Aging—A Systematic Review Focusing on Aging-Related Biomarkers. Nutrients 14, 554 (2022).

30. Ramezani, F., Herfeh, S. S. & Burke, E. The Gut Microbiota as a Mediator Linking the MIND Diet to Alzheimer’s Disease. Nutrients 18, 1445 (2026).

31. Van der Laan, M. J. & Rose, S. Targeted Learning: Causal Inference for Observational and Experimental Data. vol. 4 (Springer, 2011).

32. Thorpe, M. G., Milte, C. M., Crawford, D. & McNaughton, S. A. Education and lifestyle predict change in dietary patterns and diet quality of adults 55 years and over. Nutr J 18, 67 (2019).

33. Samieri, C., Okereke, O. I., E. Devore, E. & Grodstein, F. Long-Term Adherence to the Mediterranean Diet Is Associated with Overall Cognitive Status, but Not Cognitive Decline, in Women123. J Nutr 143, 493–499 (2013).

34. Harrington, J. M., Dahly, D. L., Fitzgerald, A. P., Gilthorpe, M. S. & Perry, I. J. Capturing changes in dietary patterns among older adults: a latent class analysis of an ageing Irish cohort. Public Health Nutr 17, 2674–2686 (2014).

35. Wortmann, H. R., Gisch, U. A., Bergmann, M. M. & Warschburger, P. Exploring the Longitudinal Stability of Food Neophilia and Dietary Quality and Their Prospective Relationship in Older Adults: A Cross-Lagged Panel Analysis. Nutrients 15, 1248 (2023).

36. Cespedes, E. M. et al. Change in Dietary Patterns and Change in Waist Circumference and DXA Trunk Fat among Post-menopausal Women. Obesity (Silver Spring) 24, 2176–2184 (2016).

37. Wang, M. et al. Alcohol consumption and epigenetic age acceleration across human adulthood. Aging (Albany NY) 15, 10938–10971 (2023).

38. Sonnega, A. et al. Cohort profile: the Health and Retirement Study (HRS). Int J Epidemiol 43, 576–585 (2014).

39. Health and Retirement Study. 2016 Venous Blood Study (VBS) (Final V3.0). 38281 Institute for Social Research, University of Michigan 10.7826/ORTM1684 (2025).

40. Lee, H. et al. Ultraprocessed Foods and the Risk of Cognitive Impairment.

41. Athey, S., Tibshirani, J. & Wager, S. Generalized random forests. Ann. Stat. 47, 1148–1178 (2019).

42. Yadlowsky, S., Fleming, S., Shah, N., Brunskill, E. & Wager, S. Evaluating Treatment Prioritization Rules via Rank-Weighted Average Treatment Effects. J Am Stat Assoc 120, 38–51 (2025).

